# Assessing Inflammatory Protein Biomarkers in COPD Subjects with and without Alpha-1 Antitrypsin Deficiency

**DOI:** 10.1101/2025.01.11.25320392

**Authors:** Matthew Moll, Brian D. Hobbs, Katherine A. Pratte, Chengyue Zhang, Auyon J. Ghosh, Russell P. Bowler, David A Lomas, Edwin K. Silverman, Dawn L. DeMeo

## Abstract

**Rationale:** Individuals homozygous for the Alpha-1 Antitrypsin (AAT) Z allele (Pi*ZZ) exhibit heterogeneity in COPD risk. COPD occurrence in non-smokers with AAT deficiency (AATD) suggests inflammatory processes may contribute to COPD risk independently of smoking. We hypothesized that inflammatory protein biomarkers in non-AATD COPD are associated with moderate-to-severe COPD in AATD individuals, after accounting for clinical factors.

**Methods:** Participants from the COPDGene (Pi*MM) and AAT Genetic Modifier Study (Pi*ZZ) were included. Proteins associated with FEV_1_/FVC were identified, adjusting for confounders and familial relatedness. Lung-specific protein-protein interaction (PPI) networks were constructed. Proteins associated with AAT augmentation therapy were identified, and drug repurposing analyses performed. A protein risk score (protRS) was developed in COPDGene and validated in AAT GMS using AUC analysis. Machine learning ranked proteomic predictors, adjusting for age, sex, and smoking history.

**Results:** Among 4,446 Pi*MM and 352 Pi*ZZ individuals, sixteen blood proteins were associated with airflow obstruction, fourteen of which were highly expressed in lung. PPI networks implicated regulation of immune system function, cytokine and interleukin signaling, and matrix metalloproteinases. Eleven proteins, including IL4R, were linked to augmentation therapy. Drug repurposing identified antibiotics, thyroid medications, hormone therapies, and antihistamines as potential AATD treatments. Adding protRS improved COPD prediction in AAT GMS (AUC 0.86 vs. 0.80, p = 0.0001). AGER was the top-ranked protein predictor of COPD.

**Conclusions:** Sixteen proteins are associated with COPD and inflammatory processes that predict airflow obstruction in AATD after accounting for age and smoking. Immune activation and inflammation are modulators of COPD risk in AATD.

## Introduction

Chronic obstructive pulmonary disease (COPD) is a major cause of morbidity and mortality worldwide^1^. A monogenic cause of COPD is severe alpha-1 antitrypsin (AAT) deficiency (AATD). AAT is encoded by the *SERPINA1* gene and is a potent inhibitor of neutrophil elastase^2^. Individuals homozygous for two Z alleles (Glu342Lys; denoted Pi*ZZ) in this gene have very low circulating serum AAT levels. AATD is associated with severe early-onset emphysema, airflow limitation, hepatic disease, and other disorders^2^. However, there is marked heterogeneity amongst individuals with Pi*ZZ with respect to the development of airflow obstruction and emphysema.

To examine factors associated with severity of lung disease amongst individuals with Pi*ZZ, the AAT Genetic Modifier Study (AAT GMS), enrolled a large cohort of index and non-index family members homozygous for the Z allele. From this cohort, cigarette smoking, male sex, asthma, pneumonia, and chronic bronchitis have previously been identified as risk factors for lower spirometry measures^3^. Serban and colleagues demonstrated that there are shared proteomic predictors of airflow obstruction and emphysema in individuals with and without Pi*ZZ, and that a protein risk score can predict emphysema^4^. However, in their analysis of 237 Pi*ZZ subjects, a lung-specific protein-protein interaction analysis of overlapping proteomic predictors with airflow obstruction was not performed, and the protein risk score was not tested in the context of a clinical risk score. Further, the proteomic platforms in this prior study were not enriched for inflammatory markers, which may offer a more global view of proteomic alterations but may also limit identification of targetable inflammatory pathways.

While smoking cessation is paramount for preventing airflow obstruction, some individuals with AATD will develop lung disease despite never smoking or quitting smoking.

Thus, there may be inflammatory processes associated with AATD leading to airflow obstruction that are independent of cigarette smoking, though this hypothesis has not been tested. Despite AATD being monogenic in etiology, the heterogeneity in disease severity and response to AAT protein replacement (hereafter, “augmentation”) therapy suggest that additional biological processes linked to AATD remain to be understood. As with other causes of COPD, inflammation could be an important driver of disease risk and severity, and leveraging a proteomic panel enriched for inflammatory protein biomarkers could identify pathogenic pathways associated with COPD and AATD.

In this study, we utilize proteomic data from the Genetic Epidemiology of COPD (COPDGene) study to train a predictive model and AAT GMS individuals with proteomic data enriched for inflammatory markers to address these issues. We hypothesized that after accounting for clinical risk factors of disease severity, there are inflammatory protein biomarkers that can predict which individuals with severe AATD will develop moderate-to-severe COPD. We additionally examined inflammatory proteins associated with AAT augmentation therapy.

## Methods

### Study populations

#### COPDGene

The Genetic Epidemiology of COPD (COPDGene) study^5^ included 10,198 non-Hispanic white (NHW) and African American (AA) individuals, 45-80 years of age with 10 or more pack-years of cigarette smoking exposure. Baseline demographic, spirometry, chest computed tomography (CT) imaging data, and whole blood samples were collected.

At the five-year follow up visit, blood samples on 5,670 individuals were collected and proteomic data was measured using SomaScan 5K (version 4.0). Further details regarding SomaScan data can be found in the Supplement. In the current analysis, we included only individuals with inferred Pi*MM based on exclusion of other genotypes, as previously reported^6^, with SomaScan and spirometry data collected at the five-year follow up visit.

#### AAT Genetic Modifiers Study

The AAT Genetic Modifier Study (AAT GMS)^3^ is a multicenter cross-sectional study of 378 European ancestry participants with severe AATD (all Pi*ZZ) in 167 families^3^. Eligible families included those with at least one sibling pair with Pi*ZZ in which both siblings were 30 years of age or greater. Questionnaire, spirometry, and whole blood were collected. Proband status was defined as the first individual in the family diagnosed with AATD.

Proteomic data were generated using the Olink Explore Inflammatory 384 panel by Olink (Waltham, MA) and preprocessed to remove outliers^7^. Data were transformed on a Log2 scale with the measurement unit on the relative NPX scale per Olink^8,9^. NPX (Normalized Protein eXpression) is a relative quantification metric used to represent protein levels detected in Olink assays. NPX is based on Proximity Extension Assay (PEA) technology, which enables sensitive, precise protein detection across a broad dynamic range. Biomarkers on this panel were chosen to represent proteins in biological pathways that most contribute to key research questions in 5 main areas: secreted proteins, organ-specific proteins, inflammatory proteins, established and ongoing drug targets, and exploratory proteins. Additional details on preparation of Olink proteomic data are in the Supplement. We included individuals with Olink proteomic and spirometry data.

## Statistical analysis

### Overview of study design

A schematic of our study design is shown in Figure 1. The study included biological characterization of proteins associated with FEV_1_/FVC and development of a proteomic predictor of FEV_1_/FVC, as well as an examination of the top proteomic predictors of airflow obstruction after accounting for clinical risk factors. For the biological characterization portion, we identified which proteins were associated with FEV_1_/FVC in both COPDGene and AAT GMS and used the replicable set of proteins to perform pathway enrichment and protein-protein interaction network analyses to gain insight into the biological meaning of our findings. We mapped these proteins to lung cell types and performed drug repurposing analysis (see below). As secondary analyses, we also examined the proteomic markers associated with AAT augmentation therapy.

**Figure 1:**
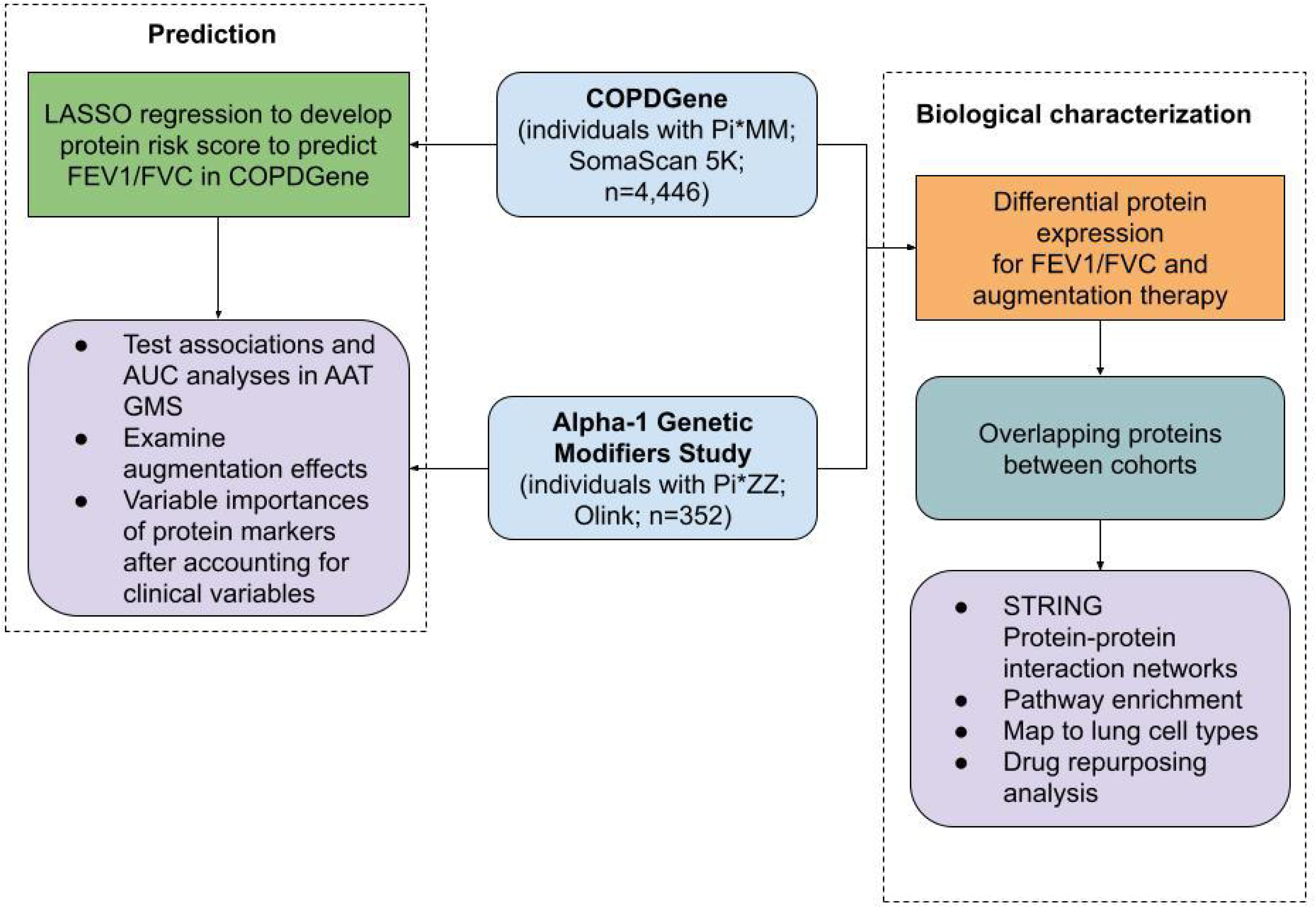
Schematic of study design. COPDGene = Genetic Epidemiology of COPD study. FEV_1_=forced expiratory volume in 1 second. FVC = forced vital capacity. LASSO = least absolute shrinkage and selection operator. Pi = alpha-1 antitrypsin protease inhibitor. AUC = area-under-the-receiver-operating-characteristic curve. STRING= search tool for the retrieval of interacting proteins.

### Biological characterization of proteomic associations with phenotypes of interest

#### Phenotypes/Outcomes of Interest

The primary phenotype or outcome of interest was FEV_1_/FVC in both cohorts. In the AAT GMS cohort, we also tested a range of secondary associations of interest including with AAT augmentation therapy administration, C-Reactive Protein (CRP) (measured separately from Olink), bronchodilator responsiveness (BDR), immunoglobulin E (IgE), and FEV_1_ % predicted.

Given the right skew of FEV_1_/FVC, we used rank-normalized FEV_1_/FVC for all analyses. SomaScan and Olink proteins, CRP, and IgE were log2-transformed prior to analysis.

#### Biological characterization of proteins associated with FEV1/FVC

We performed differential protein expression analysis in COPDGene and AAT GMS for each phenotype of interest. We first limited to proteins present in both the COPDGene and AAT GMS datasets based on overlapping UniProt identifiers (272 proteins). In COPDGene, we performed analyses using multiple linear regressions, adjusting for potential confounders, including age, sex, self-identified race, current smoking status, pack-years of smoking, and study center. In AAT GMS, we applied linear mixed effects models utilizing the OlinkAnalyze R package (https://github.com/Olink-Proteomics/OlinkRPackage) olink_lmer function. We estimated effect sizes and confidence intervals with the lmerTest R package lmer or glmer functions for continuous and binary outcomes, respectively, considering family relatedness (i.e., identifiers) as random intercepts. We additionally adjusted models for age, sex, pack-years of smoking, pack-years of smoking squared, ever smoking status, and proband status as fixed effects. As a sensitivity analysis, we additionally adjusted models for augmentation therapy, which can alter proteomic associations^4^. In both cohorts, we considered Benjamini-Hochberg^10^ p-values less than 0.05 to be significant. We applied this same approach to identify proteomic markers associated with augmentation therapy.

We focused remaining biological characterization analyses on FEV_1_/FVC. We compared the effect sizes and directions of each protein associated with FEV_1_/FVC in each cohort to identify a list of replicable protein biomarkers. In AAT GMS, we used Pearson correlation coefficients to examine the correlation of phenotypes and FEV_1_/FVC-associated proteins with each other and constructed correlation plots with ggcorrplot.

We mapped the list of replicable proteins associated with FEV1/FVC to a human lung single cell atlas^11^ and used these proteins to build a protein-protein interaction (PPI) network (https://string-db.org/). The rationale for using the human cell atlas via Cell-X-Gene was to leverage its broader tissue-specific and cell-specific data coverage compared to other databases, such as GTEx, in providing sufficient cellular representation for the selected proteins. We then performed pathway enrichment and Enrichr^12–14^ drug repurposing analyses based on this network. Details regarding these analyses are in the Supplement.

### Prediction of spirometric severity in individuals with Pi*ZZ

Details regarding the development of clinical and protein risk scores for FEV1/FVC are in the Supplement.

#### Testing of the protein risk score

We first tested the association of the protRS with multiple outcomes in COPDGene and AAT GMS using multivariable linear regression models. Outcomes tested are detailed in the Supplement.

After performing association analyses, we then performed area-under-the-receiver-operating-characteristic-curve (AUC) analyses to evaluate the predictive performance of the clinical risk score (CRS), protRS, and both CRS and protRS together for COPD case-control status (Global Initiative for Chronic Obstructive Lung Disease (GOLD) 2-4 versus normal spirometry). We compared AUCs considering DeLong p-values^15^ below 0.05 as indicating a significant difference. We also split the protRS into tertiles and examined the odds of having moderate-to-severe COPD for individuals in the second and third compared to the first tertile.

To identify the relative importances of proteins that predict moderate-to-severe COPD in individuals with AATD after accounting for clinical risk factors, we obtained residuals for linear regression models of each protRS protein with clinical factors (protein ∼ age + sex + pack-years of smoking). Using the residuals of these models as inputs, we developed a random forest model, which allows for modeling of non-linear relationships and provides variable importance measures. The random forest model was trained in the AAT GMS with FEV_1_/FVC as the outcome with 500 trees and 5 variables tested at each split. Variable importances were based on changes in mean squared error (MSE) – that is, when a protein is removed from the model, there is a resulting increase in the MSE; the greater the increase in MSE, the more important the variable.

## Results

### Characteristics of study participants

Characteristics of study participants are shown in Table 1. We included 352 Pi*ZZ individuals from the Alpha-1 Genetic Modifiers Study (AAT GMS) and 4,446 Pi*MM subjects from the Genetic Epidemiology of COPD (COPDGene) study. Compared to COPDGene, AAT GMS participants were all non-Hispanic white and were more likely to be younger, female, have fewer pack-years of smoking history, and lower FEV_1_ and FEV_1_/FVC. Within AAT GMS, the correlation between spirometry phenotypes was strong, but limited between spirometry and other traits (Figure S1)

**Table 1.** Characteristics of study participants in the Alpha-1 Genetic Modifier Study (AAT GMS; all Pi*ZZ) and Genetic Epidemiology of COPD study (COPDGene; limited to Pi*MM). FEV_1_ = forced expiratory volume in 1 second. FEV_1_/FVC = FEV_1_/forced vital capacity. LAA = low attenuation area. HU = Hounsfield units. Perc15 = 15th percentile of lung density histogram on inspiratory CT scans. WA % = wall area percent. Pi10 = square root of wall area of a hypothetical internal perimeter of 10 mm.

| *Characteristic* | *AAT GMS* | *COPDGene* |
| --- | --- | --- |
| N | 352 | 4446 |
| age (mean (SD)) | 51.42 (9.03) | 65.53 (8.63) |
| sex (No. (%), female) | 195 (55.4) | 2165 (48.7) |
| race (No. (%), African American) | 0 (0.0) | 1380 (31.0) |
| Pack-years of smoking (mean (SD)) | 13.34 (15.93) | 44.28 (24.28) |
| Current smoking status (No. (%)) | 12 (3.4) | 1712 (38.5) |
| Ever smoking status (No. (%)) | 236 (67.0) | 4446 (100.0) |
| FEV<sub>1</sub> (mean (SD)) | 2.03 (1.18) | 2.16 (0.87) |
| FEV<sub>1</sub>/FVC (mean (SD)) | 0.54 (0.21) | 0.66 (0.15) |
| % LAA < -950 HU (mean (SD)) | NA | 5.78 (9.13) |
| Perc15 (mean (SD)) | NA | 82.06 (28.72) |
| WA% (mean (SD)) | NA | 49.83 (8.39) |
| Pi10 (mean (SD)) | NA | 2.26 (0.58) |
| augmentation therapy (No. (%)) | 161 (45.7) | NA |

### Proteins associated with FEV_1_/FVC and other phenotypes in individuals with Pi*MM and Pi*MZ

Differential protein expression results for all phenotypes in the AAT GMS are shown in Table S1. We identified 78 proteins significantly associated with FEV_1_/FVC in COPDGene individuals with Pi*MM and 67 proteins significantly associated with FEV_1_/FVC in AAT GMS individuals with Pi*ZZ; AAT GMS effects were similar after adjusting for augmentation therapy (Table S2). We found 20 proteins associated with FEV_1_ that were not associated with FEV_1_/FVC (Table S3). Comparing results across cohorts, there were 16 overlapping significantly differentially expressed proteins based on UniProt IDs (Table 2). We observed concordant directions of effects for each protein except for ADCYAP1 (AAT GMS: ß = 0.156, COPDGene: ß = -0.0935). Given the different proteomic platforms, direct comparisons of effect sizes cannot be interpreted, only directions of effects. Examination of the heatmap in Figure S2 demonstrates that LY9 and CD48 are highly correlated (r ≥ 0.8), but the other 14 proteins are less highly correlated.

**Table 2:** Proteins significantly associated with FEV_1_/FVC in both the Alpha-1 Genetic Modifier Study and COPDGene. See Table 1 for abbreviations. In the Alpha-1 Genetic Modifier Study (AAT GMS), models were adjusted for age, sex, pack-years of smoking, pack-years of smoking squared, ever smoking status, and proband status. In COPDGene, models were adjusted for age, sex, race, height, current smoking status, pack-years of smoking, and study center. The table is arranged by the adjusted p-values for AAT GMS. *ADCYAP1 is another name for PACAP

| UniProt ID | HGNC | AAT GMS |  | COPDGene |  |
| --- | --- | --- | --- | --- | --- |
|  |  | *beta (95% CI)* | *adj. p-value* | *beta (95% CI)* | *adj. p-value* |
| P12544 | GZMA | 0.167 (0.105 to 0.228) | 1.60E-05 | 0.118 (0.0255 to 0.21) | 0.044 |
| Q15109 | AGER | 0.379 (0.225 to 0.534) | 2.20E-05 | 0.123 (0.0757 to 0.17) | 8.20E-06 |
| P21709 | EPHA1 | 0.38 (0.21 to 0.55) | 2.00E-04 | 0.196 (0.129 to 0.262) | 5.00E-07 |
| P29460 | IL12B | 0.217 (0.107 to 0.328) | 0.00036 | 0.14 (0.0622 to 0.218) | 0.0039 |
| Q9HBG7 | LY9 | 0.444 (0.245 to 0.644) | 0.00036 | 0.199 (0.118 to 0.28) | 2.70E-05 |
| P09326 | CD48 | 0.422 (0.212 to 0.631) | 0.00066 | 0.211 (0.127 to 0.296) | 2.10E-05 |
| Q99983 | OMD | 0.225 (0.113 to 0.337) | 0.0027 | 0.13 (0.0549 to 0.206) | 0.0056 |
| Q15661 | TPSAB1 | 0.221 (0.0963 to 0.346) | 0.0031 | 0.186 (0.117 to 0.255) | 3.50E-06 |
| P43489 | TNFRSF4 | 0.256 (0.0867 to 0.425) | 0.0099 | 0.166 (0.0339 to 0.297) | 0.049 |
| P48023 | FASLG | 0.278 (0.107 to 0.45) | 0.01 | 0.0918 (0.0238 to 0.16) | 0.031 |
| Q99435 | NELL2 | 0.337 (0.142 to 0.532) | 0.01 | 0.273 (0.0995 to 0.446) | 0.012 |
| Q9Y6N7 | ROBO1 | 0.323 (0.126 to 0.519) | 0.012 | 0.253 (0.149 to 0.357) | 3.00E-05 |
| O00241 | SIRPB1 | 0.178 (0.0366 to 0.319) | 0.026 | 0.0678 (0.0162 to 0.119) | 0.037 |
| Q14005 | IL16 | 0.112 (0.0237 to 0.201) | 0.036 | 0.286 (0.184 to 0.388) | 1.30E-06 |
| Q8WU39 | ADCYAP1 | 0.156 (0.0377 to 0.274) | 0.036 | -0.0935 (-0.161 to -0.0257) | 0.028 |
| P15260 | IFNGR1 | 0.397 (0.0982 to 0.696) | 0.037 | 0.159 (0.0565 to 0.262) | 0.013 |

We then mapped these 16 proteins to the Cell-X-Gene human lung single cell atlas to identify those with gene expression levels in the top quartile for each lung cell type (Figure 2, refer to Supplementary Methods). This analysis revealed that all but two proteins, IL12B and ADCYAP1, were likely to have high expression levels in lung tissue, and the other 14 proteins were thus selected for subsequent network analysis. Using these 14 proteins, we constructed a protein-protein interaction network (Figure 3, Table S4) and performed MCL clustering analysis, which defined three clusters (Table S5). Notably, the first cluster implicates processes related to regulation of cytotoxicity and immunoregulatory interactions between lymphoid and non-lymphoid cells, while the third cluster implicates activation of matrix metalloproteinases. STRING-based Reactome pathway enrichment analyses implicate alterations in immune system function, cytokine signaling, and interleukin signaling (Table S6).

**Figure 2.**
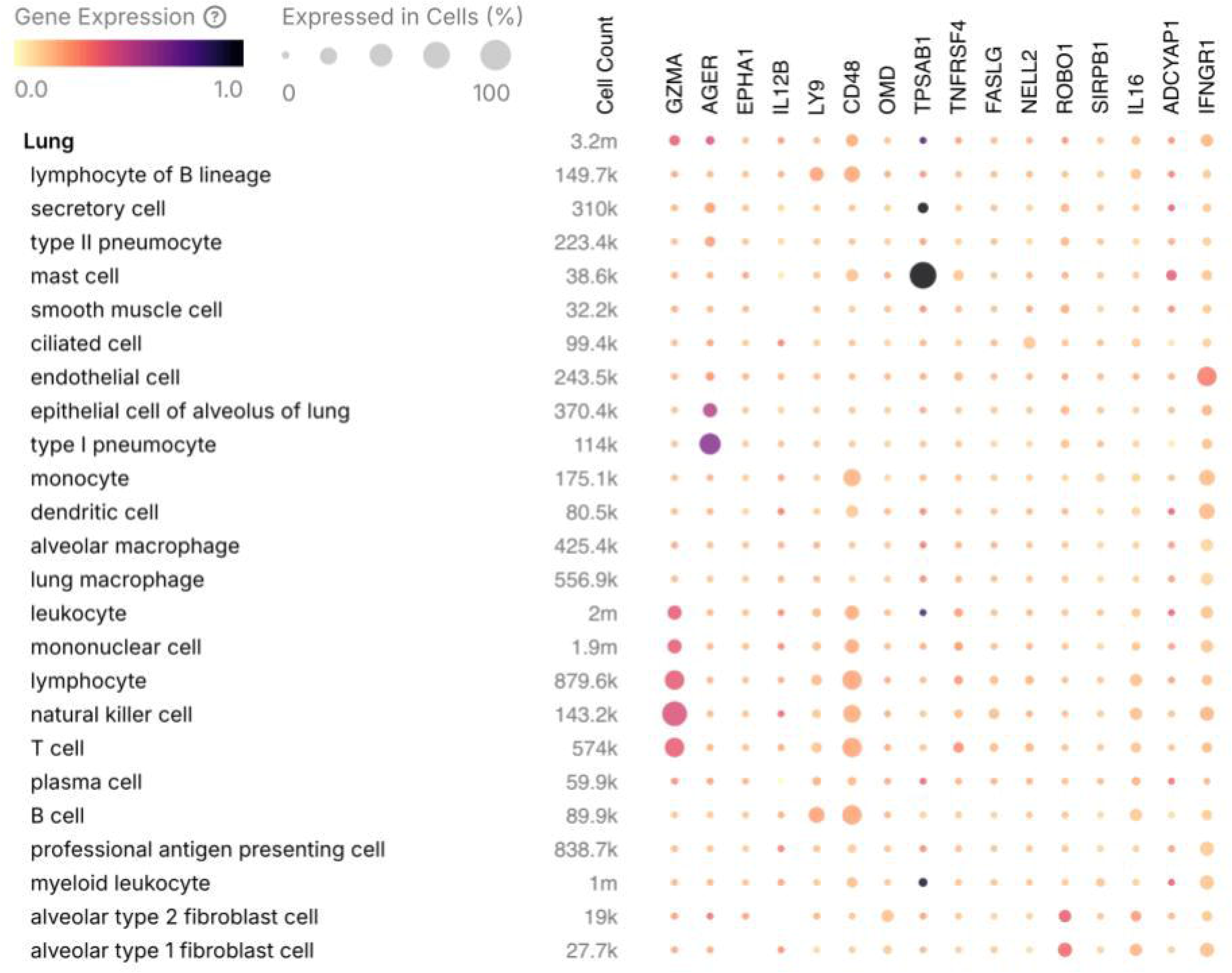
The top 16 proteins associated with FEV_1_/FVC in both Alpha-1 Genetic Modifiers Study and COPDGene were mapped to the human lung single cell atlas (https://cellxgene.cziscience.com/gene-expression). *ADCYAP1 is another name for PACAP.

**Figure 3.**
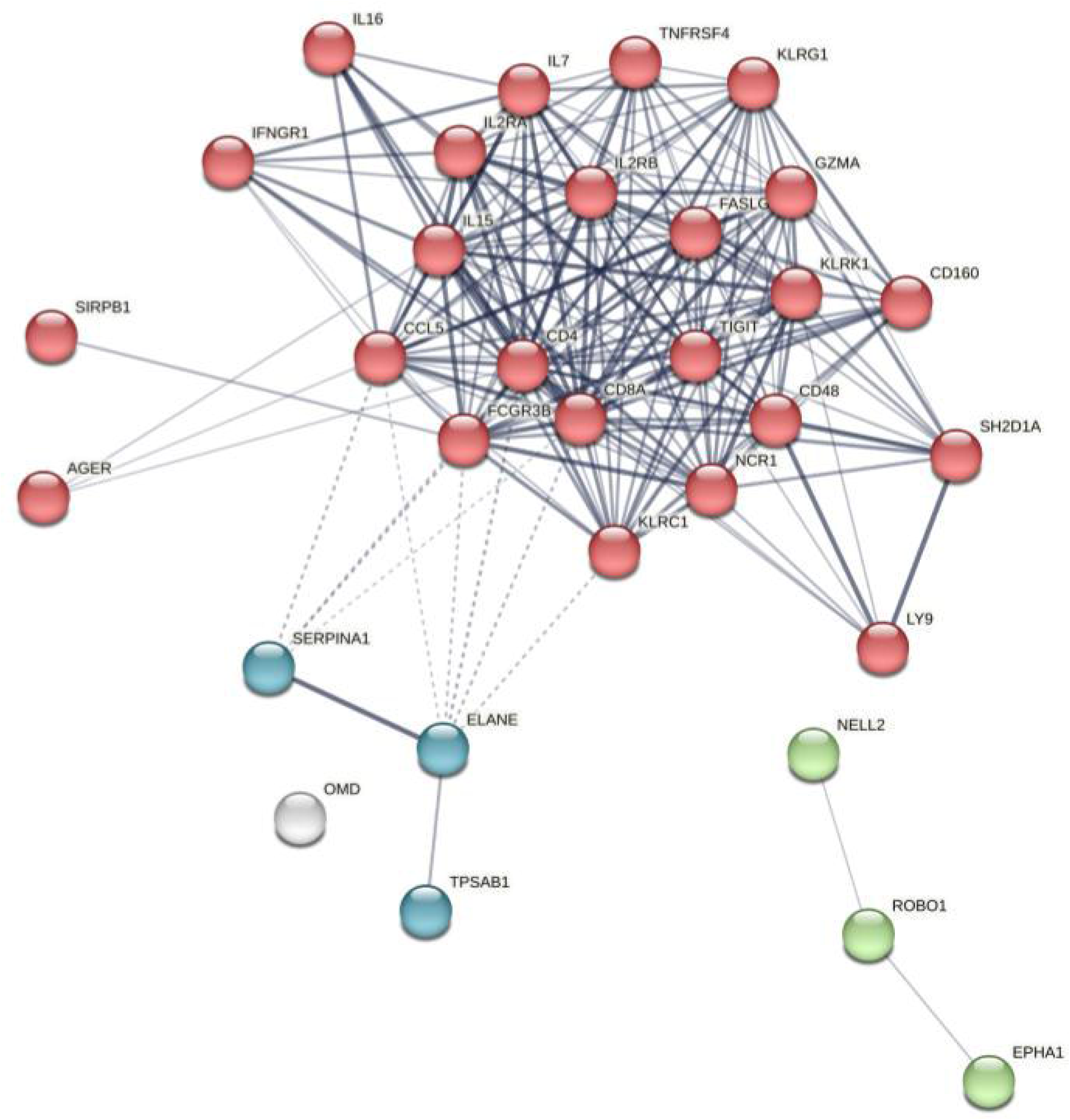
STRING network built from top 14 proteins associated with FEV1/FVC in both Alpha-1 Genetic Modifiers Study and COPDGene with high expression in lung cells (top quartile of expression). Medium confidence interactions were included (>0.4). Edge thickness indicates level of confidence. Edges with 10 interactors in the first shell and 5 in the second shell were permitted. MCL clustering was performed with and inflation factor of 3 to define clusters, which are shown in different colors.

Using the full set of proteins in the STRING network (Table S4), we performed enrichment-based drug-repurposing analyses (Table 3). We identified 12 drug repurposing candidates of interest, though only methimazole was significant after adjusting for multiple statistical comparisons. Drug candidates of interest included antihistamines, antivirals, and thyroid medications. Steroids were identified, which are currently used for COPD. Several immunosuppressive medications (e.g. decitabine) and hormone-related therapies (e.g. flutamide) were also identified but may not have appropriate side effect profiles for use in AATD patients; these exploratory analyses point to potentially targetable pathways for further investigation.

**Table 3.** Proteins in the top quartile of expression in lung cells were mapped to the human protein-protein interactome using STRING. The full set of proteins comprising the STRING network (Table S4) were used as inputs into Enrichr to query the Multi-marker Analysis of GenoMic Annotation (MAGMA) drugs and diseases database, which identified drugs that would target enriched pathways represented by the proteins associated with FEV_1_/FVC in both the Alpha-1 Genetic Modifiers Study and COPDGene.

| *term* | *p-value* | *q-value* |
| --- | --- | --- |
| Methimazole | 0.0004 | 0.0189 |
| Sirolimus | 0.0035 | 0.0633 |
| Progesterone | 0.0059 | 0.0787 |
| Cetirizine | 0.0077 | 0.0787 |
| Nafamostat | 0.0169 | 0.0787 |
| Decitabine | 0.0184 | 0.0787 |
| Didanosine | 0.0200 | 0.0787 |
| Methylprednisolone | 0.0245 | 0.0787 |
| Propylthiouracil | 0.0260 | 0.0787 |
| Flutamide | 0.0306 | 0.0787 |
| Cladribine | 0.0321 | 0.0787 |
| Prednisolone | 0.0396 | 0.0821 |

#### Proteomic alterations associated with augmentation therapy

As AAT augmentation therapy may have anti-inflammatory effects and/or contribute additional proteins to plasma, we examined the proteomic associations with the use of augmentation therapy. We found 11 proteins significantly associated with augmentation therapy (Table S7) and only EPHA1 and AGER were present in the list of replicable proteins associated with FEV_1_/FVC (Table 2). Using these proteins in Enrichr drug repurposing analyses, we identified 5 candidates, including a macrolide antibiotic and fibrates (Table S8).

#### A protein risk score for FEV_1_/FVC predicts spirometry severity in individuals Pi*ZZ

Having identified shared proteomic associations with FEV_1_/FVC in both COPDGene and the AAT GMS, we then constructed a protein risk score (protRS) that can predict moderate-to-severe COPD status in COPDGene participants (Table S9 and S10); details of the protRS development, including hyperparameter tuning (Figure S3) and performance measures are in the Supplement. We calculated the protRS in AAT GMS and observed that it was associated with FEV_1_/FVC (Figure S4), COPD case-control status and augmentation therapy administration (Figure S5) in unadjusted analyses. In multivariable linear mixed effects analysis, the protRS was associated with FEV_1_ and FEV_1_/FVC, but not CRP or IgE levels (Table S11). A one standard deviation increase in the protRS was associated with an adjusted odds ratio of 2.8 (95% CI: 1.79 to 4.42, p = 0.000008) for moderate-to-severe COPD.

We then assessed the predictive value of the protRS in the context of known clinical risk factors. First, we developed a clinical risk score to predict FEV_1_/FVC in COPDGene using age, sex, and pack-years of smoking. We tested the predictive performance of these models in the AAT GMS (Figure 4A). In AUC analyses, we found that a clinical risk score (AUC 0.8) and the protRS similarly predicted COPD case-control status (AUC 0.81). Combining the clinical risk score and the protRS improved the AUC to 0.86 (p [AUC Combined model vs. AUC Clinical model] = 1E- 04 (Figure 4A).

**Figure 4.**
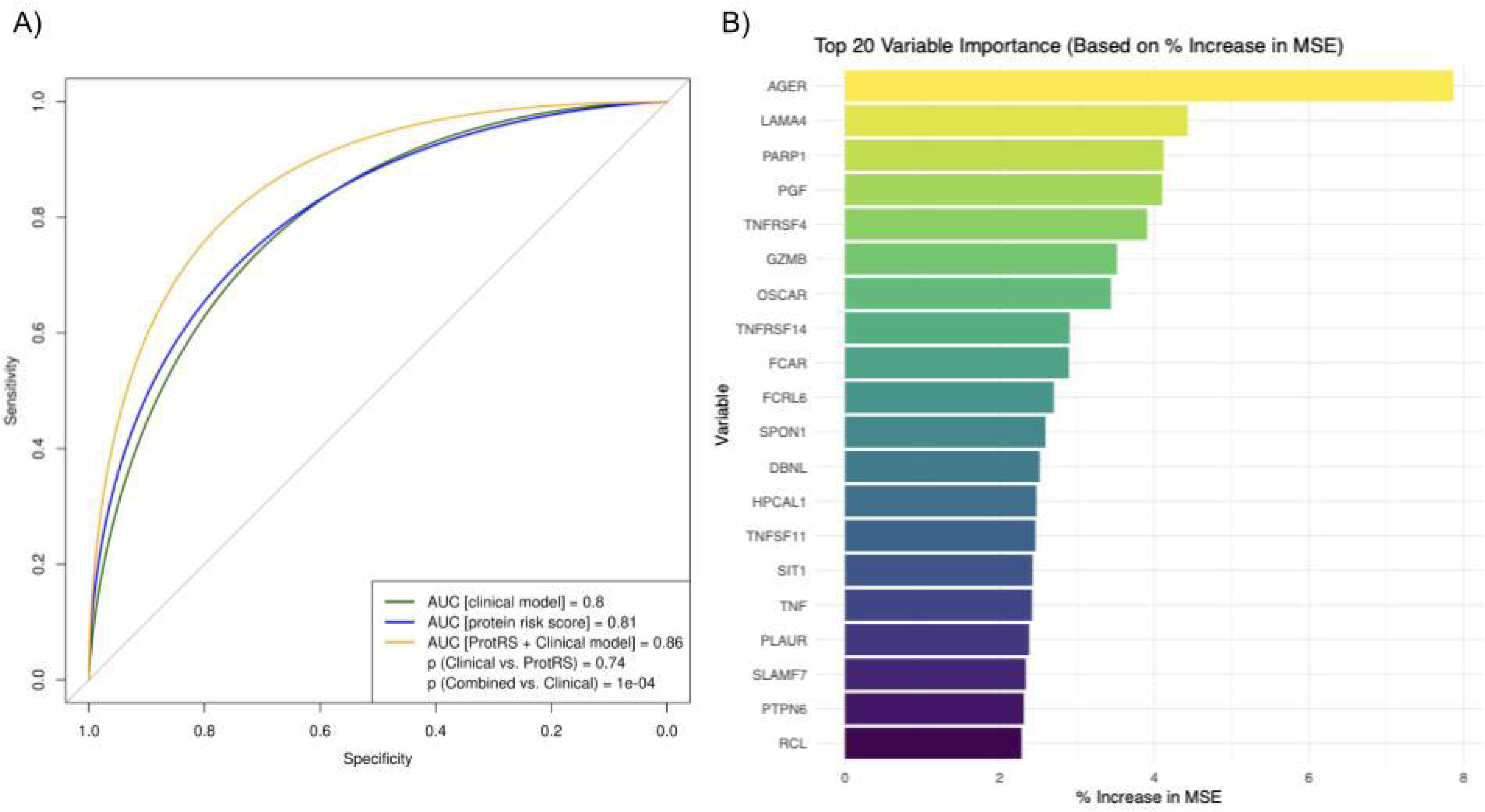
A) Receiver-operating-characteristic curves and area-under-the-curve measures for models trained in COPDGene and tested in the Alpha-1 Genetic Modifiers Study. The clinical model included age, sex, and pack-years of smoking. ProtRS=protein risk score. ‘Combined’ indicates the combination of clinical variables and the protRS. B) Random forest model-based variable importance measures for proteins in the protein risk score after adjusting for age, sex, and pack-years of smoking.

To identify the most important inflammatory proteins predictive of airflow obstruction in individuals with AATD, we first adjusted the protein NPX values for age, sex, and pack-years of smoking. We then used the residuals of these protein regression models to train a random forest model in the AAT GMS. After removing the effects of the clinical factors, the random forest model explained 6.53% of the variance in FEV_1_/FVC, and the top 20 most important variables are shown in Figure 4B, with the most important protein predictor being AGER.

## Discussion

In this study of over 4,000 individuals with Pi*MM who smoked and 352 individuals with Pi*ZZ, we identified 16 proteins measured in plasma that were associated with airflow obstruction in both cohorts. Fourteen of these proteins were highly expressed in lung. Using protein-protein interaction network analyses, we found that these proteins are involved in regulating immune system function, cytokine signaling, interleukin signaling, and matrix metalloproteinases. We identified drug repurposing candidates that included antihistamines, antivirals, hormone therapies, and thyroid medications. We also identified 11 proteins associated with AAT augmentation therapy. We then developed a protein risk score (protRS) that predicts moderate-to-severe COPD within individuals with Pi*ZZ and was additive to clinical risk factors. Finally, we used machine learning to identify inflammatory proteins that predict airflow obstruction within individuals with Pi*ZZ, after accounting for clinical risk factors. These results lend insight into the inflammatory processes contributing to airflow obstruction in individuals with Pi*ZZ beyond age, sex, and cigarette smoking exposure, and identify potential therapeutic targets and drug repurposing candidates for future research.

Our results extend the work of Serban and colleagues who previously demonstrated shared proteomic markers between individuals with Pi*MM and Pi*ZZ with respect to airflow obstruction and emphysema^4^. The authors also developed a protein risk score that predicted emphysema in individuals with Pi*ZZ. Our study expands upon these findings in the following important ways: we (1) used an AATD cohort that was not examined in the Serban et al. study^4^, larger in size, and focused on inflammatory protein biomarkers, (2) performed a lung-specific PPI network analysis of the replicable proteins associated with FEV_1_/FVC, (3) performed drug repurposing analyses, (4) directly examined proteins associated with AAT augmentation therapy, (5) tested the performance of the protRS in the context of a clinical risk score, (6) used machine learning to identify inflammatory drivers of airflow obstruction after accounting for clinical factors.

Our findings suggest that individuals with AATD have ongoing inflammation that contributes to airflow obstruction regardless of cigarette smoking exposure. We observed that clinical risk factors, including but not limited to pack-years of smoking, and proteins in the protein risk score explained ∼25% of the variance in FEV_1_/FVC, while proteins adjusted for clinical variables explained 6.5% of the variance. In this analysis, *AGER*, a highly replicable COPD GWAS locus and biomarker for emphysema in non-AATD subjects, was ranked by random forest as the most important protein predictor. The rs2070600 variant encodes a missense polymorphism in the *AGER* gene and its association with COPD and emphysema have been replicated in multiple GWASs^16,17^. The protein product of *AGER* is soluble receptor for advanced glycation end-products (s-RAGE), a multi-ligand transmembrane receptor expressed in lung and other tissues that is implicated in several inflammatory diseases. Lower levels are associated with more emphysema and greater COPD risk^18,19^. Thus, understanding the role of AGER in both non-AATD and AATD emphysema and COPD as a mechanistic target and biomarker is an important area for additional investigation.

We identified 16 inflammatory proteins associated with FEV_1_/FVC in both COPDGene and the AAT GMS. All 16 of these proteins were reported to be associated with FEV_1_/FVC by Serban et al.^4^. Fourteen of these proteins (excluding IL12B and ADCYAP1) were highly expressed in lung, and a network-based enrichment analysis implicated alterations in immunoregulatory interactions between lymphoid and non-lymphoid cells, changes in cytokine and interleukin signaling, and regulation of matrix metalloproteinases. These processes have been previously implicated in COPD pathogenesis. B-cells and lymphoid follicle density in the lung are associated with disease severity and emphysema^20,21^. AAT has been shown to attenuate cytokine and interleukin inflammatory responses^22^, including in the context of infections^23^.

We demonstrated that a 126-protein risk score improved predictive capacity for COPD affection status in individuals with Pi*ZZ above a clinical risk score. A protein panel of this size, enriched for inflammatory biomarkers, likely has high translatability across cohorts and proteomic assay platforms. Our results also confirm that the proteomic determinants of disease severity in non-AATD and AATD COPD are at least partially shared, as previously reported^4^, and suggest that blood-based biomarkers developed in individuals without AATD may be useful in individuals with AATD.

We further identified 11 proteins associated with AAT augmentation therapy which implicate alterations in immune function with changes in IL-4, IL-17, and IL-24; whether these proteins can be leveraged as additional therapeutic targets and/or predict response to augmentation therapy requires further study. Two proteins overlapped with those proteins associated with FEV_1_/FVC (AGER, EPHA1), which suggests that augmentation therapy may alter airflow obstruction risk through these proteins.

As augmentation therapy is effective yet far from curative, we performed drug repurposing analyses which identified several drug repurposing candidates. Several antibiotics and antiviral agents were identified, which seems plausible given that most COPD exacerbations are related to bacterial and viral infections. Indeed, a successful drug repurposing agent for COPD exacerbations is the macrolide antibiotic azithromycin^24^ and roxithromycin was identified as a candidate based on augmentation-associated proteins. Currently, we do not prescribe antivirals for COPD exacerbations, but our results suggest this approach may be worth consideration. Some of the drug repurposing candidates may be targeting comorbid conditions such as allergic rhinitis (antihistamines). While thyroid medications were identified as potential drug candidates, there is limited literature to support this finding beyond a previously reported association between thyroid dysfunction and COPD^25^. Notably, methimazole is used to treat hyperthyroidism, raising questions about whether the direction of effect aligns with the intended therapeutic outcome. Drugs within the same class may have the same primary mechanism of action yet variable off-target effects, so our findings should not be interpreted solely at the drug class level. While our results are intriguing, to consider using these agents in AATD patients, careful pharmacoepidemiologic or randomized controlled trials need to be performed.

Nonetheless, our results highlight the need to apply additional drug repurposing approaches to future datasets and consider the best way to test drug candidates beyond AAT protein replacement therapy.

Strengths of this study include leveraging a proteomic platform enriched for inflammatory biomarkers and machine learning to identify biological processes related to airflow obstruction in a large cohort of individuals with Pi*ZZ after accounting for age and smoking, utilizing a network-based approach to understand these biological processes, examining proteomic associations with additional outcomes (e.g., CRP, IgE, augmentation therapy), and performing drug repurposing analyses.

Limitations include that we did not have lung proteomic data from individuals with AATD, and we cannot decipher whether protein associations are causing or caused by disease- associated phenotypes. However, we did map our protein associations to a human lung single- cell atlas to focus on proteins expressed in lung tissue. Prospective validation of the protRS would be necessary before clinical use and an additional replication of the protRS in another AATD population would lend support to a prospective trial of such a biomarker. While we found proteins associations with FEV_1_/FVC that replicated in terms of significance and direction of effect, effect sizes cannot be inferred due to the cross-platform (SomaScan and Olink) nature of this study. Further research into cross-platform proteomic analyses are needed. The drug repurposing results yielded many intriguing trends, but enrichment results did not pass multiple comparison testing at a strict statistical threshold for the primary analysis, though it did in the analysis of augmentation therapy-associated proteins; these results might be driven by the sparsity of our network. We also cannot infer directionality with respect the how drug repurposing candidates may alter disease risk, and this issue needs to be addressed before designing validation studies. Having concomitant single cell data with proteomic and drug repurposing analyses could help identify testable and targetable pathways within specific cell types.

In conclusion, we identified 14 lung-expressed proteins associated with COPD severity and identified inflammatory proteins and pathways associated with airflow obstruction in individuals with AATD that persist after adjusting for the effects of age and smoking. Further, we identified drug repurposing candidates and proteins associated with AAT augmentation therapy and developed a protein risk score that improves prediction of COPD affection status when added to clinical factors. Further validation and investigation of our findings can lead to an improved understanding of the pathogenesis of airflow obstruction and potential therapeutic strategies for people with severe Alpha-1 Antitrypsin deficiency.

## Data Availability

All data produced in the present study are available upon reasonable request to the authors. COPDGene data are available through dbGaP.

## Author contributions

Study Design: Matthew Moll, Brian D. Hobbs, Dawn L. DeMeo

Acquisition, analysis, or interpretation of the data: Matthew Moll, Chengyue Zhang, Edwin K. Silverman, Katherine Pratte, Russell Bowler, Brian D. Hobbs, David Lomas, Dawn L. DeMeo

Critical revision of the manuscript for important intellectual content: All authors

Statistical analysis: Matthew Moll, Brian D. Hobbs, Katherine Pratte.

Obtained funding: Dawn L. DeMeo

## References

1. Safiri, S. et al. Burden of chronic obstructive pulmonary disease and its attributable risk factors in 204 countries and territories, 1990-2019: results from the Global Burden of Disease Study 2019. BMJ **378**, e069679 (2022).

2. Silverman, E. K. & Sandhaus, R. A. Clinical practice. Alpha1-antitrypsin deficiency. The New England journal of medicine 360, 2749–57 (2009).

3. Demeo, D. L. et al. Determinants of airflow obstruction in severe alpha-1-antitrypsin deficiency. Thorax 62, 806–813 (2007).

4. Serban, K. A. et al. Unique and shared systemic biomarkers for emphysema in Alpha-1 Antitrypsin deficiency and chronic obstructive pulmonary disease. EBioMedicine 84, 104262 (2022).

5. Regan, E. A. et al. Genetic epidemiology of COPD (COPDGene) study design. COPD 7, 32–43 (2010).

6. Ghosh, A. J. et al. Alpha-1 Antitrypsin MZ Heterozygosity Is an Endotype of Chronic Obstructive Pulmonary Disease. Am J Respir Crit Care Med 205, 313–323 (2022).

7. Assarsson, E. et al. Homogenous 96-plex PEA immunoassay exhibiting high sensitivity, specificity, and excellent scalability. PLoS One 9, e95192 (2014).

8. Wik, L. et al. Proximity Extension Assay in Combination with Next-Generation Sequencing for High-throughput Proteome-wide Analysis. Mol Cell Proteomics 20, 100168 (2021).

9. Bowman, W. S. et al. Proteomic biomarkers of progressive fibrosing interstitial lung disease: a multicentre cohort analysis. Lancet Respir Med 10, 593–602 (2022).

10. Benjamini, Y. & Hochberg, Y. Controlling the False Discovery Rate: A Practical and Powerful Approach to Multiple Testing. Journal of the Royal Statistical Society. Series B (Methodological) 57, 289–300 (1995).

11. Megill, C. et al. cellxgene: a performant, scalable exploration platform for high dimensional sparse matrices. 2021.04.05.438318 Preprint at 10.1101/2021.04.05.438318 (2021).

12. Chen, E. Y. et al. Enrichr: interactive and collaborative HTML5 gene list enrichment analysis tool. BMC Bioinformatics 14, 128 (2013).

13. Kuleshov, M. V. et al. Enrichr: a comprehensive gene set enrichment analysis web server 2016 update. Nucleic Acids Res 44, W90–97 (2016).

14. Xie, Z. et al. Gene Set Knowledge Discovery with Enrichr. Current Protocols 1, e90 (2021).

15. DeLong, E. R., DeLong, D. M. & Clarke-Pearson, D. L. Comparing the areas under two or more correlated receiver operating characteristic curves: a nonparametric approach. Biometrics 44, 837–45 (1988).

16. Sakornsakolpat, P. et al. Genetic landscape of chronic obstructive pulmonary disease identifies heterogeneous cell-type and phenotype associations. Nature genetics 51, 494–505 (2019).

17. Shrine, N. et al. Multi-ancestry genome-wide association analyses improve resolution of genes and pathways influencing lung function and chronic obstructive pulmonary disease risk. Nat Genet 55, 410–422 (2023).

18. Sin, S., Lim, M., Kim, J., Bak, S. H. & Kim, W. J. Association between plasma sRAGE and emphysema according to the genotypes of AGER gene. BMC Pulm Med 22, 58 (2022).

19. Pratte, K. A. et al. Soluble receptor for advanced glycation end products (sRAGE) as a biomarker of COPD. Respiratory Research 22, 127 (2021).

20. Rojas-Quintero, J. et al. Spatial Transcriptomics Resolve an Emphysema-Specific Lymphoid Follicle B Cell Signature in Chronic Obstructive Pulmonary Disease. Am J Respir Crit Care Med 209, 48–58 (2024).

21. Faner, R. et al. Network Analysis of Lung Transcriptomics Reveals a Distinct B-Cell Signature in Emphysema. American journal of respiratory and critical care medicine 193, 1242–53 (2016).

22. Campos, M. A. & Geraghty, P. Cytokine Regulation by Alpha-1 Antitrypsin Therapy: A Pathway Analysis of a Pilot Clinical Trial. Am J Respir Cell Mol Biol 66, 697–700 (2022).

23. Pott, G. B., Chan, E. D., Dinarello, C. A. & Shapiro, L. α-1-Antitrypsin is an endogenous inhibitor of proinflammatory cytokine production in whole blood. J Leukoc Biol 85, 886–895 (2009).

24. Albert, R. K. et al. Azithromycin for prevention of exacerbations of COPD. N Engl J Med 365, 689–698 (2011).

25. Chaudhary, S. C. et al. Prevalence of thyroid dysfunction in chronic obstructive pulmonary disease patients in a tertiary care center in North India. Journal of Family Medicine and Primary Care 7, 584 (2018).

