## Supplementary appendix for "Assessing Inflammatory Protein Biomarkers in COPD Subjects with and without Alpha-1 Antitrypsin Deficiency"

### Funding and Acknowledgements

**COPDGene Phase 3 Grant Support and Disclaimer**

The project described was supported by Award Number U01 HL089897 and Award Number U01 HL089856 from the National Heart, Lung, and Blood Institute. The content is solely the responsibility of the authors and does not necessarily represent the official views of the National Heart, Lung, and Blood Institute or the National Institutes of Health.

**COPD Foundation Funding**

COPDGene is also supported by the COPD Foundation through contributions made to an Industry Advisory Board that has included AstraZeneca, Bayer Pharmaceuticals, Boehringer- Ingelheim, Genentech, GlaxoSmithKline, Novartis, Pfizer, and Sunovion.

**COPDGene® Investigators – Core Units**

*Administrative Center*: James D. Crapo, MD (PI); Edwin K. Silverman, MD, PhD (PI); Barry J. Make, MD; Elizabeth A. Regan, MD, PhD

*Genetic Analysis Center*: Terri H. Beaty, PhD; Peter J. Castaldi, MD, MSc; Michael H. Cho, MD, MPH; Dawn L. DeMeo, MD, MPH; Adel El Boueiz, MD, MMSc; Marilyn G. Foreman, MD, MS; Auyon Ghosh, MD; Lystra P. Hayden, MD, MMSc; Craig P. Hersh, MD, MPH; Jacqueline Hetmanski, MS; Brian D. Hobbs, MD, MMSc; John E. Hokanson, MPH, PhD; Wonji Kim, PhD; Nan Laird, PhD; Christoph Lange, PhD; Sharon M. Lutz, PhD; Merry-Lynn McDonald, PhD; Dmitry Prokopenko, PhD; Matthew Moll, MD, MPH; Jarrett Morrow, PhD; Dandi Qiao, PhD; Elizabeth A. Regan, MD, PhD; Aabida Saferali, PhD; Phuwanat Sakornsakolpat, MD; Edwin K. Silverman, MD, PhD; Emily S. Wan, MD; Jeong Yun, MD, MPH

*Imaging Center*: Juan Pablo Centeno; Jean-Paul Charbonnier, PhD; Harvey O. Coxson, PhD; Craig J. Galban, PhD; MeiLan K. Han, MD, MS; Eric A. Hoffman, Stephen Humphries, PhD; Francine L. Jacobson, MD, MPH; Philip F. Judy, PhD; Ella A. Kazerooni, MD; Alex Kluiber; David A. Lynch, MB; Pietro Nardelli, PhD; John D. Newell, Jr., MD; Aleena Notary; Andrea Oh, MD; Elizabeth A. Regan, MD, PhD; James C. Ross, PhD; Raul San Jose Estepar, PhD; Joyce Schroeder, MD; Jered Sieren; Berend C. Stoel, PhD; Juerg Tschirren, PhD; Edwin Van Beek, MD, PhD; Bram van Ginneken, PhD; Eva van Rikxoort, PhD; Gonzalo Vegas Sanchez- Ferrero, PhD; Lucas Veitel; George R. Washko, MD; Carla G. Wilson, MS;

*PFT QA Center, Salt Lake City, UT*: Robert Jensen, PhD

*Data Coordinating Center and Biostatistics*, *National Jewish Health, Denver, CO*: Douglas Everett, PhD; Jim Crooks, PhD; Katherine Pratte, PhD; Matt Strand, PhD; Carla G. Wilson, MS

*Epidemiology Core*, *University of Colorado Anschutz Medical Campus, Aurora, CO*: John E. Hokanson, MPH, PhD; Erin Austin, PhD; Gregory Kinney, MPH, PhD; Sharon M. Lutz, PhD; Kendra A. Young, PhD

*Mortality Adjudication Core:* Surya P. Bhatt, MD; Jessica Bon, MD; Alejandro A. Diaz, MD, MPH; MeiLan K. Han, MD, MS; Barry Make, MD; Susan Murray, ScD; Elizabeth Regan, MD; Xavier Soler, MD; Carla G. Wilson, MS

*Biomarker Core*: Russell P. Bowler, MD, PhD; Katerina Kechris, PhD; Farnoush Banaei- Kashani, PhD

**COPDGene® Investigators – Clinical Centers**

*Ann Arbor VA:* Jeffrey L. Curtis, MD; Perry G. Pernicano, MD

*Baylor College of Medicine, Houston, TX*: Nicola Hanania, MD, MS; Mustafa Atik, MD; Aladin Boriek, PhD; Kalpatha Guntupalli, MD; Elizabeth Guy, MD; Amit Parulekar, MD;

*Brigham and Women’s Hospital, Boston, MA*: Dawn L. DeMeo, MD, MPH; Craig Hersh, MD, MPH; Francine L. Jacobson, MD, MPH; George Washko, MD

*Columbia University, New York, NY*: R. Graham Barr, MD, DrPH; John Austin, MD; Belinda D’Souza, MD; Byron Thomashow, MD

*Duke University Medical Center, Durham, NC*: Neil MacIntyre, Jr., MD; H. Page McAdams, MD; Lacey Washington, MD

*HealthPartners Research Institute, Minneapolis, MN*: Charlene McEvoy, MD, MPH; Joseph Tashjian, MD

*Johns Hopkins University, Baltimore, MD*: Robert Wise, MD; Robert Brown, MD; Nadia N. Hansel, MD, MPH; Karen Horton, MD; Allison Lambert, MD, MHS; Nirupama Putcha, MD, MHS

*Lundquist Institute for Biomedical Innovation at Harbor UCLA Medical Center, Torrance, CA*: Richard Casaburi, PhD, MD; Alessandra Adami, PhD; Matthew Budoff, MD; Hans Fischer, MD; Janos Porszasz, MD, PhD; Harry Rossiter, PhD; William Stringer, MD

*Michael E. DeBakey VAMC, Houston*, *TX*: Amir Sharafkhaneh, MD, PhD; Charlie Lan, DO *Minneapolis VA:* Christine Wendt, MD; Brian Bell, MD; Ken M. Kunisaki, MD, MS

*Morehouse School of Medicine, Atlanta, GA*: Eric L. Flenaugh, MD; Hirut Gebrekristos, PhD; Mario Ponce, MD; Silanath Terpenning, MD; Gloria Westney, MD, MS

*National Jewish Health, Denver, CO*: Russell Bowler, MD, PhD; David A. Lynch, MB *Reliant Medical Group, Worcester, MA*: Richard Rosiello, MD; David Pace, MD

*Temple University, Philadelphia, PA:* Gerard Criner, MD; David Ciccolella, MD; Francis Cordova, MD; Chandra Dass, MD; Gilbert D’Alonzo, DO; Parag Desai, MD; Michael Jacobs, PharmD; Steven Kelsen, MD, PhD; Victor Kim, MD; A. James Mamary, MD; Nathaniel Marchetti, DO; Aditi Satti, MD; Kartik Shenoy, MD; Robert M. Steiner, MD; Alex Swift, MD; Irene Swift, MD; Maria Elena Vega-Sanchez, MD

*University of Alabama, Birmingham, AL:* Mark Dransfield, MD; William Bailey, MD; Surya P. Bhatt, MD; Anand Iyer, MD; Hrudaya Nath, MD; J. Michael Wells, MD

*University of California, San Diego, CA*: Douglas Conrad, MD; Xavier Soler, MD, PhD; Andrew Yen, MD

*University of Iowa, Iowa City, IA*: Alejandro P. Comellas, MD; Karin F. Hoth, PhD; John Newell, Jr., MD; Brad Thompson, MD

*University of Michigan, Ann Arbor, MI:* MeiLan K. Han, MD MS; Ella Kazerooni, MD MS; Wassim Labaki, MD MS; Craig Galban, PhD; Dharshan Vummidi, MD

*University of Minnesota, Minneapolis, MN*: Joanne Billings, MD; Abbie Begnaud, MD; Tadashi Allen, MD

*University of Pittsburgh, Pittsburgh, PA*: Frank Sciurba, MD; Jessica Bon, MD; Divay Chandra, MD, MSc; Joel Weissfeld, MD, MPH

*University of Texas Health, San Antonio, San Antonio, TX*: Antonio Anzueto, MD; Sandra Adams, MD; Diego Maselli-Caceres, MD; Mario E. Ruiz, MD; Harjinder Singh

### Supplementary Methods

##### COPDGene SomaScan data

Plate hybridization, median signal normalization, and plate scaling and calibration of SOMAmers were performed to control for variability across array signals, inter-run variability, inter-assay variation between analytes and batch differences between plates. Median normalization to a reference using adaptive normalization by maximum likelihood is applied within SOMAmer dilution group to quality control replicates and on individual samples to remove edge effect and technical variance. Data were log2-transformed prior to statistical analysis. Further details regarding preparation of SomaScan data has been previously published^1^.

##### Alpha-1 Antitrypsin Genetic Modifier Study Olink Data

The Olink Explore Inflammatory 384 panel was employed to assess a comprehensive array of inflammatory biomarkers in Alpha-1 Antitrypsin Genetic Modifier Study (AAT GMS) participants. This panel utilizes proximity extension assay (PEA) technology, which enables simultaneous quantification of 384 protein biomarkers from a minute volume of sample. Briefly, the assay involves a dual-antibody recognition approach where pairs of antibodies coupled to unique DNA oligonucleotides bind to target proteins in the sample. Upon binding, the DNA oligonucleotides are brought into proximity, allowing DNA polymerization and subsequent quantification via real-time PCR. The resulting double-stranded DNA sequences are specific to each antigen and are amplified using P5 and P7 Illumina adaptors along with sample indexing. These amplified targets were quantified through next-generation sequencing. To correct for systematic errors, protein estimates were normalized across different batches and log2-transformed to achieve a normal distribution, which was visually confirmed using histograms. No imputation was performed for the missing data.

#### Network and drug repurposing analysis

We mapped the list of replicable proteins associated with FEV_1_/FVC to a human lung single cell atlas (<https://cellxgene.cziscience.com/gene-expression>) and filtered for genes with expression levels in the upper quartile in at least one lung cell type. The selected lung cell types represent critical immune cells and various organ compartments. As SERPINA1 and ELANE (neutrophil elastase) were not on the Olink panel, but are central to AATD pathogenesis, we added these proteins to the list. We then mapped these proteins to the human protein-protein interactome using STRING ([www.string-db.org](http://www.string-db.org)). We only included high confidence interactions (>0.4). We permitted 10 interactors in the first shell and 5 in the second shell. MCL clustering was performed with and inflation factor of 3 to define clusters. To identify drug repurposing candidates, we used all of the proteins within the STRING network to perform enrichment-based drug repurposing analysis using Enrichr^2–4^ ([www.enrichr.org](http://www.enrichr.org)) and referencing the Multi-marker Analysis of GenoMic Annotation (MAGMA) database of drugs and diseases^5^. We considered drugs with p-values below 0.05 to be of interest, and q-value less than 0.05 to be significant.

#### Development of clinical and protein risk scores to predict FEV_1_/FVC

We developed a protein risk score (protRS) to predict FEV_1_/FVC using COPDGene participants and tested the protRS performance in AGMS. To develop the score, we used least absolute shrinkage and selection operator (LASSO) models, training to an outcome of FEV_1_/FVC, and tuned hyperparameters using 10-fold cross-validation to optimize the mean squared error. As inputs, we compared two models: (1) the overlapping differentially expressed proteins between COPDGene and AGMS, and (2) the entire set of 272 shared proteomic markers available in both datasets. Using the same LASSO parameters, we also developed a clinical risk score (CRS) in COPDGene using age, sex, and pack-years of smoking. We referenced the transparent reporting of a multivariable prediction model for individual prognosis or diagnosis (TRIPOD) reporting standards^6^ to ensure transparent reporting of our prediction model.

#### Testing of the protein risk score

In COPDGene, we examined the following outcomes: FEV_1_, FEV_1_/FVC, computed tomography (CT) measures of quantitative emphysema (quantitative emphysema on inspiratory CT scans (% LAA < -950 HU)^7^, 15th percentile of lung density histogram on inspiratory CT scans (Perc15)^8^) and of airway thickening (wall area percent (WA%)^7^ and square root of wall area of a hypothetical internal perimeter of 10 mm (Pi10)^9^); in AGMS, we examined the following outcomes: FEV_1_, FEV_1_/FVC, IgE, CRP, and moderate-to-severe COPD case-control status (GOLD 2-4 vs. normal spirometry). Models were adjusted for the covariates as detailed above for each cohort.

### Supplementary Results

#### Characteristics of Study Participants

The standard deviation in FEV_1_ was larger in AGMS (1.18 L) compared to COPDGene (0.87 L). 161 (46%) of AGMS participants reported ever receiving augmentation therapy. We examined the correlation of AGMS phenotypes with each other, and found that cross-sectional spirometry measures (FEV_1_, FEV_1_ % predicted, and FEV_1_/FVC) were highly correlated to each other, while bronchodilator responsiveness, CRP, and IgE were weakly correlated to other phenotypes (Figure S1).

#### Development of a protein risk score

We trained and compared two LASSO regression models in COPDGene: (1) using the 16 overlapping proteins as inputs, and (2) using all available proteins as inputs. The first model included 11 proteins and had an adjusted R^2^ value of 0.0316 while the latter model included 126 proteins (lambda = 0.00965; Figure S3) and had an adjusted R^2^ of 0.274. Therefore, we carried forward the second risk score as the protRS for our remaining analyses. Proteins and weights in the risk score are shown in Table S9. In COPDGene, the protRS was associated with FEV_1_/FVC as well as markers of emphysema and airway wall thickness (Table S10).

### Supplementary Figures

Figure S1: Correlation amongst phenotypes in the Alpha-1 Genetic Modifier Study. Pearson correlation coefficients are displayed in tiles with corresponding color intensities as indicated.

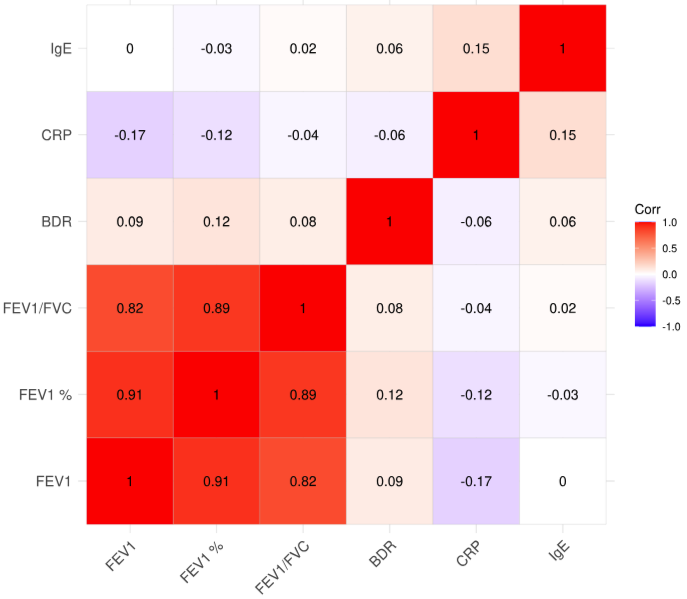

Figure S2: Correlation matrix of the 16 replicable proteins significantly associated with FEV1/FVC within the Alpha-1 Genetic Modifier Study. Pearson correlation coefficients are displayed in tiles with corresponding color intensities as indicated.

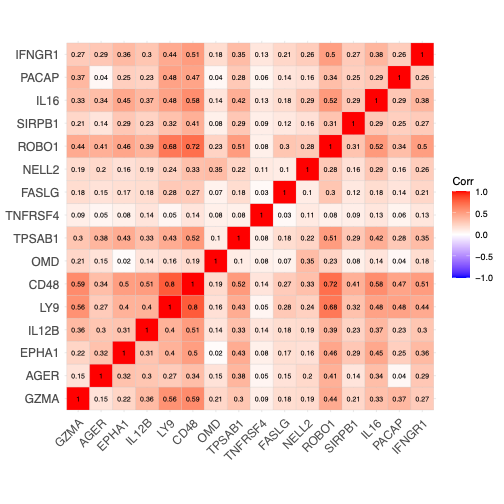

Figure S3: Cross-validation plot showing the minimum lambda based on LASSO regression of protein expression values with FEV_1_/FVC in COPDGene. The top axis indicates the number of features at each lambda value (*x-axis*).

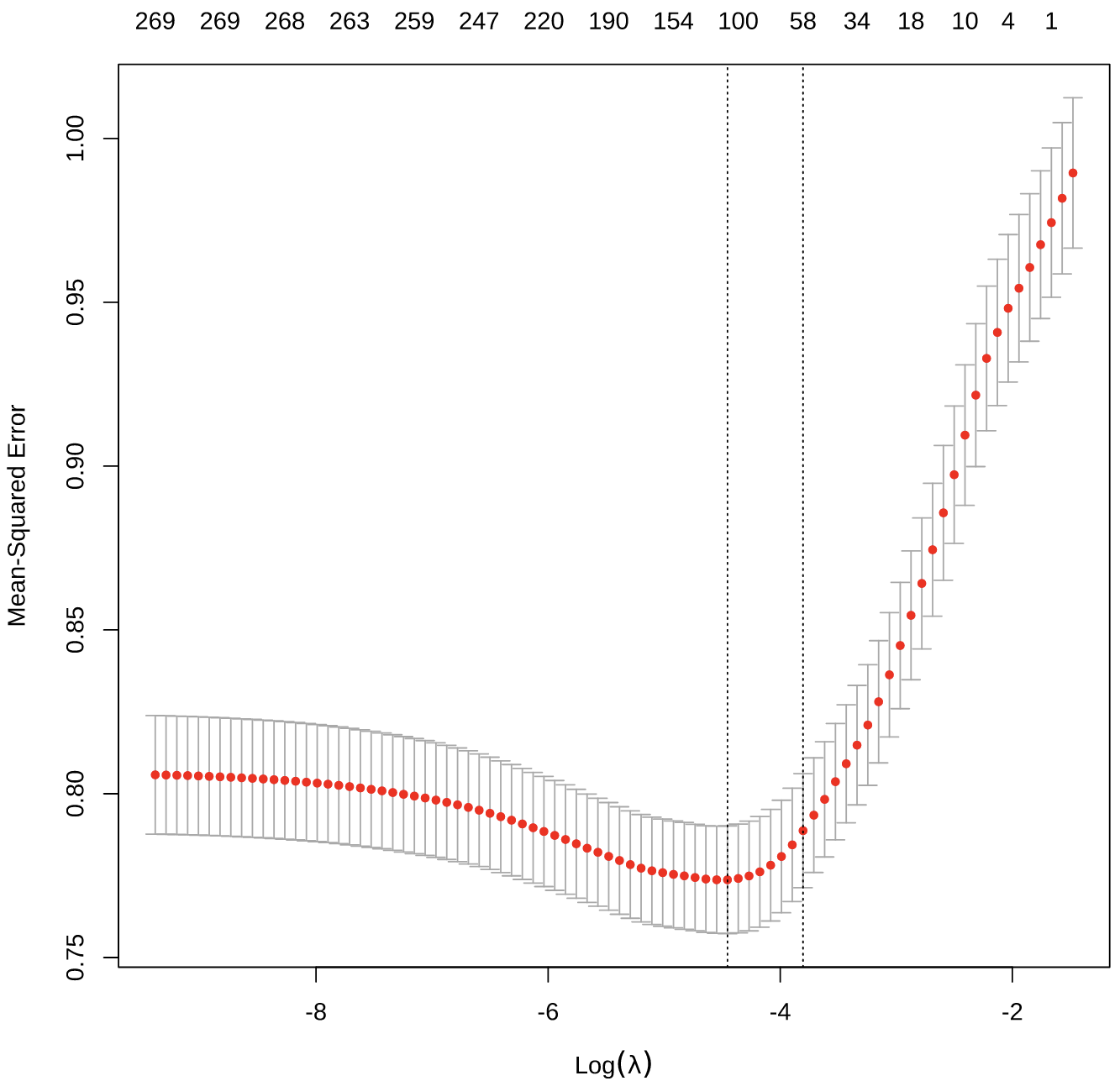

Figure S4: Scatter plot demonstrating the association between the protein risk score and FEV_1_/FVC in the Alpha-1 Genetic Modifiers Study.

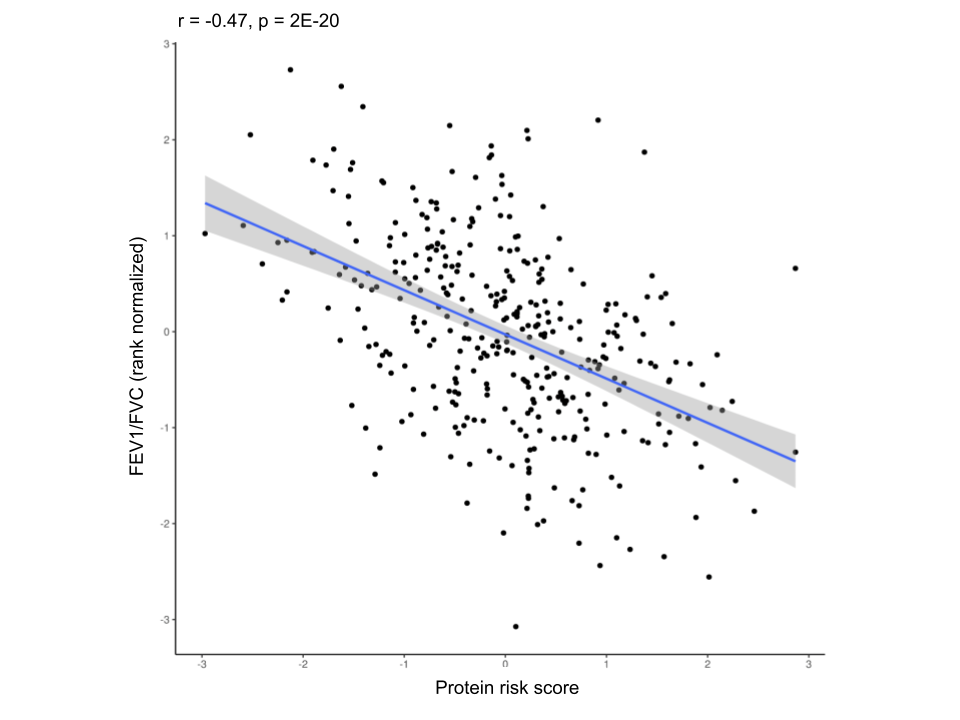

Figure S5: Violin and boxplots demonstrating the univariable association between the protein risk score developed in COPDGene and A) moderate-to-severe COPD (GOLD 2-4), and B) Augmentation therapy in the Alpha-1 Genetic Modifiers study. The boxplots indicate the median and interquartile range of the data.

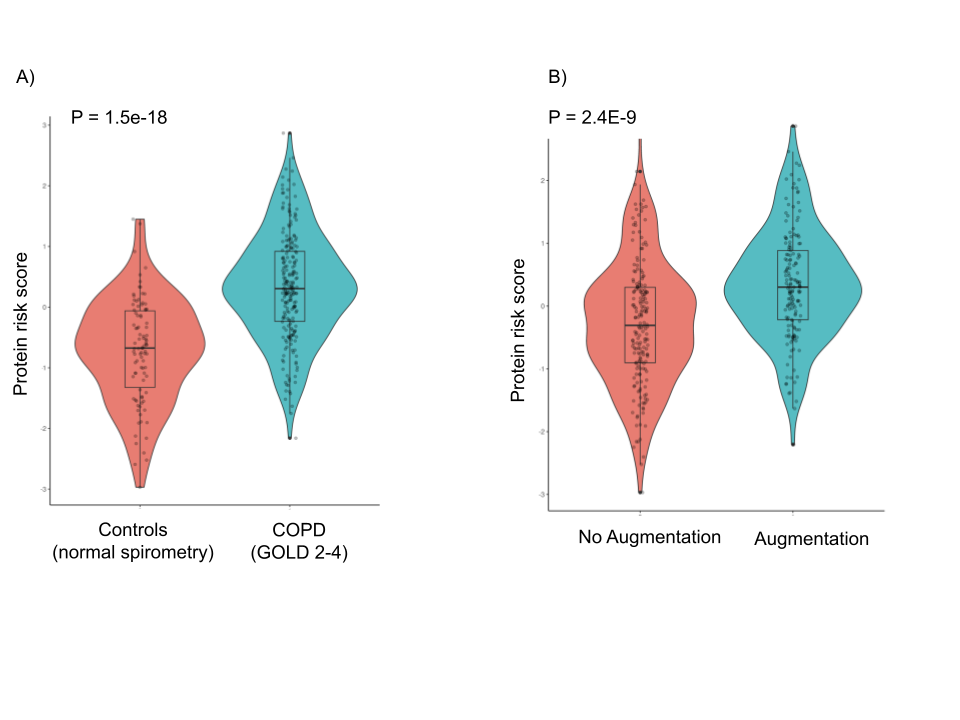

### Supplementary Tables

See SuppTables_aat_proteomics_MattMoll_freeze_11.08.2024_v2.xlsx

### Supplementary Discussion

LY9, GZMA, and CD48 were within the largest network-based cluster. These proteins have been implicated in non-AATD and AATD COPD patients. In non-AATD COPD patients, LY9 expression has previously been reported to be downregulated in COPD^10,11^. Granzyme A has been reported to be associated with COPD^12^ and is part of a family of serine proteases implicated in COPD risk. For example, Granzyme B (GzmB) is secreted by regulatory cells which can suppress CD8+ T cells, suggesting a protective role in emphysema^13^. CD48, involved in the activation of immune cells such as T cells, antigen-presenting cells, and granulocytes^14^, has been linked with nonallergic asthma^15^. In AATD individuals, a weighted gene correlation expression analysis in the bronchoalveolar lavage fluid identified clusters associated with disease severity, and these clusters included LY9, interferon genes, and GZMA^16^. Taken together, our results highlight the role of T cells and immune activation on disease severity in AATD individuals.

IFNGR1 expression is increased in COPD populations, enhances LPS signaling in alveolar macrophages of COPD patients^17^, and contributes to viral clearance the lungs^18^. TPSAB1, which encodes tryptase, is upregulated in eosinophilic COPD^19^ but was associated with higher FEV_1_/FVC in our study, suggesting that lower levels may select for non-eosinophilic COPD individuals within AATD individuals. To our knowledge, these proteins have not been investigated with respect to AATD and may represent important biomarkers and/or therapeutic targets.

### References

1.	Candia, J. *et al.* Assessment of Variability in the SOMAscan Assay. *Sci. Rep.* **7**, 14248 (2017).

2.	Chen, E. Y. *et al.* Enrichr: interactive and collaborative HTML5 gene list enrichment analysis tool. *BMC Bioinformatics* **14**, 128 (2013).

3.	Kuleshov, M. V. *et al.* Enrichr: a comprehensive gene set enrichment analysis web server 2016 update. *Nucleic Acids Res.* **44**, W90-97 (2016).

4.	Xie, Z. *et al.* Gene Set Knowledge Discovery with Enrichr. *Curr. Protoc.* **1**, e90 (2021).

5.	de Leeuw, C. A., Mooij, J. M., Heskes, T. & Posthuma, D. MAGMA: generalized gene-set analysis of GWAS data. *PLoS Comput. Biol.* **11**, e1004219 (2015).

6.	Collins, G. S., Reitsma, J. B., Altman, D. G. & Moons, K. G. M. Transparent reporting of a multivariable prediction model for individual prognosis or diagnosis (TRIPOD): the TRIPOD statement. *BMJ* **350**, g7594 (2015).

7.	Han, M. K. *et al.* Chronic Obstructive Pulmonary Disease Exacerbations in the COPDGene Study: Associated Radiologic Phenotypes. *Radiology* **261**, 274–282 (2011).

8.	Parr, D. G., Sevenoaks, M., Deng, C. Q., Stoel, B. C. & Stockley, R. A. Detection of emphysema progression in alpha 1-antitrypsin deficiency using CT densitometry; Methodological advances. *Respir. Res.* **9**, 1–8 (2008).

9.	Van Tho, N. *et al.* A mixed phenotype of airway wall thickening and emphysema is associated with dyspnea and hospitalization for chronic obstructive pulmonary disease. *Ann. Am. Thorac. Soc.* **12**, 988–996 (2015).

10.	Wu, X., Sun, X., Chen, C., Bai, C. & Wang, X. Dynamic gene expressions of peripheral blood mononuclear cells in patients with acute exacerbation of chronic obstructive pulmonary disease: a preliminary study. *Crit. Care* **18**, 508 (2014).

11.	Zhang, J. *et al.* Bioinformatics analyses of the pathogenesis and new biomarkers of chronic obstructive pulmonary disease. *Medicine (Baltimore)* **100**, e27737 (2021).

12.	Zhao, Y. *et al.* Identification of Macrophage Polarization-Related Genes as Biomarkers of Chronic Obstructive Pulmonary Disease Based on Bioinformatics Analyses. *BioMed Res. Int.* **2021**, e9921012 (2021).

13.	Kim, W.-D. *et al.* The Role of Granzyme B Containing Cells in the Progression of Chronic Obstructive Pulmonary Disease. *Tuberc. Respir. Dis.* **83**, S25–S33 (2020).

14.	McArdel, S. L., Terhorst, C. & Sharpe, A. H. Roles of CD48 in regulating immunity and tolerance. *Clin. Immunol. Orlando Fla* **164**, 10–20 (2016).

15.	Breuer, O. *et al.* Evaluation of Soluble CD48 Levels in Patients with Allergic and Nonallergic Asthma in Relation to Markers of Type 2 and Non-Type 2 Immunity: An Observational Study. *J. Immunol. Res.* **2018**, 4236263 (2018).

16.	Chu, J. *et al.* Gene Co-expression Networks Reveal Novel Molecular Endotypes in Alpha-1 Antitrypsin Deficiency. *Thorax* **76**, 134–143 (2021).

17.	Southworth, T. *et al.* IFN-γ synergistically enhances LPS signalling in alveolar macrophages from COPD patients and controls by corticosteroid-resistant STAT1 activation. *Br. J. Pharmacol.* **166**, 2070–2083 (2012).

18.	Xu, F. *et al.* The molecular and cellular mechanisms associated with the destruction of terminal bronchioles in COPD. *Eur. Respir. J.* **59**, (2022).

19.	Higham, A., Dungwa, J., Pham, T., McCrae, C. & Singh, D. Increased mast cell activation in eosinophilic chronic obstructive pulmonary disease. *Clin. Transl. Immunol.* **11**, e1417 (2022).
